# Designing Interprofessional Education in Mechanical Ventilation: A Study of Learning Needs and Gaps

**DOI:** 10.1101/2025.09.14.25335731

**Authors:** Hiroshi Tajima, Hajime Kasai, Kenichiro Takeda, Takao Utsumi, Yutaka Furukawa, Izumi Usui, Takuji Suzuki, Ikuko Sakai, Shoichi Ito

**Affiliations:** Department of Medical Education, Graduate School of Medicine, Chiba University, Chiba, Japan; Health Professional Development Centre, Chiba University Hospital, Chiba, Japan; Department of Respirology, Graduate School of Medicine, Chiba University, Chiba, Japan; Practical Pharmacy, Graduate School and Faculty of Pharmaceutical Science, Chiba University, Chiba, Japan; Department of Clinical Engineering Centre, Chiba University Hospital, Chiba, Japan

**Keywords:** Respiratory Therapy, Interprofessional Relations, Education, Medical, Needs Assessment, Curriculum Development

## Abstract

Mechanical ventilation (MV) management is a complex clinical strategy that requires interdisciplinary collaboration. The aim of this study was to evaluate the status of MV-related learning, educational needs, perceived facilitators, and barriers across multiple healthcare professions in a university hospital. A cross-sectional survey was conducted among medical doctors, nurses, clinical engineers, pharmacists, and physical therapists. The quantitative items evaluated prior experience, understanding of terminology, confidence, motivation, and preferred learning formats. Free-text responses were analysed to identify common and profession-specific learning factors. In total, 119 professionals completed the survey. Most participants had MV-related clinical experience (80.6%) and prior education (84.0%) primarily through on-the-job training and in-person lectures. Clinical experience was positively associated with higher motivation to learn (mean 4.1 vs. 3.2, p < 0.01), while motivation showed a modest decline with increasing years since licensure (p = 0.01). Across all professional groups, interest in MV education was high (mean > 4.0, across 28 items), especially in foundational knowledge, clinical decision-making, and interprofessional collaboration. Preferences for learning formats varied by profession: medical doctors and clinical engineers tended to favour simulation-based learning, whereas pharmacists and physical therapists showed a preference for asynchronous on-demand formats. Content analysis demonstrated five common facilitators: experiential learning opportunities, favourable workplace conditions, access to structured educational resources, intrinsic motivation, and organisational support. Conversely, barriers showed profession-specific patterns; excessive workload was frequently reported by nurses and physical therapists (p = 0.01), while pharmacists cited low motivation more commonly (p = 0.02). Given shared facilitators and distinct profession-specific barriers, MV education should adopt a hybrid instructional model that integrates a common interprofessional core with targeted modules tailored to each discipline’s clinical roles and learning preferences. This approach may enhance learner engagement, facilitate the practical application of MV knowledge, and strengthen interdisciplinary collaboration, thereby contributing to pandemic preparedness and improved patient outcomes.

## Introduction

Mechanical ventilators (MVs) are life-sustaining medical devices designed to supplement or replace pulmonary function [1]. They play critical roles in supporting ventilation and delivering oxygen in diverse clinical situations, such as severe respiratory failure, trauma, and postoperative care [2]. Effective MV management requires multidisciplinary knowledge and skills, including respiratory physiology, initiation and weaning protocols, device operation, and interprofessional communication across numerous clinical roles [3].

The Coronavirus disease 2019 (COVID-19) pandemic, which emerged in late 2019, has led to a dramatic surge in patients with pneumonia-induced respiratory failure requiring mechanical ventilation [4]. Healthcare systems are overwhelming globally [5], and many healthcare workers, including those without formal intensive care training, are pressed into frontline role [6]. Given the recurring nature of global respiratory outbreaks (e.g. severe acute respiratory syndrome [SARS] in 2002 and Middle East Respiratory Syndrome [MERS] in 2012) [7], the ability to rapidly mobilise interprofessional teams equipped with MV competencies is crucial for maintaining healthcare system resilience in future pandemics [8].

However, MV education remains fragmented and insufficiently formalised. MV education lacks standardisation and relies heavily on institutional discretion and individual mentorship. Previous studies have underscored the widespread uncertainty regarding learners’ preparedness for MV and the lack of consensus on optimal teaching and assessment methods [9]. Learners often report low satisfaction because of time constraints and the absence of clear learning goals [10]. These challenges underscore the urgent need to establish structured and inclusive MV education programs.

Moreover, MV management is inherently interprofessional and requires collaboration among many kinds of professionals [11–13]. When conducting interprofessional collaborative practice, interprofessional competency frameworks emphasise core domains such as role contribution, relationship facilitation, reflection, and understanding others [14] [15]. However, the educational needs and barriers specific to each profession including shared challenges across roles remain underexplored in the context of MV education.

To develop structured and inclusive MV education programs, it is essential to understand current learning conditions, profession-specific needs, and shared challenges among healthcare professionals. Specifically, this study addresses the following research questions: 1) How do the learning status and motivation for MV management differ among healthcare professionals, and what commonalities exist across them? and 2) What are the characteristics of profession-specific learning needs and shared educational priorities across all professions? 3) What facilitating and inhibiting factors affect MV-related learning, and how do they vary or align across different professional roles?

By clarifying both common ground and profession-specific distinctions, this study aims to inform the design of practical and sustainable interprofessional education (IPE) programs that contribute to pandemic preparedness and long-term team capacity building.

## Materials and Methods

### 1. Participants

All healthcare professionals working at Chiba University Hospital during the survey period were invited to participate. There were no restrictions based on profession during the recruitment process, and those who completed the full questionnaire were included in the analysis. Incomplete responses and those from individuals not engaged in patient care were excluded.

### 2. Questionnaire Survey Content

We conducted a cross-sectional online survey between October 1st and December 28th, 2024, using Google Forms® to assess the MV learning status, needs, and perceptions of healthcare professionals at Chiba University Hospital.

Table 1 outlines the questionnaire items that addressed the participants’ characteristics (age, sex, and years since licensure; Q1–Q3), prior learning opportunities, understanding of MV-related terminology, and clinical experience (Q4–Q6). Items also evaluated self-reported confidence in MV management and motivation to learn (Q7–Q8) as well as preferences regarding educational formats and interest in specific learning content (Q9–Q11). Finally, the open-ended questions explored the perceived facilitators and barriers to MV-related learning (Q12 and Q13). The questionnaire was developed based on a review of the existing literature on MV education and interprofessional learning needs [16–18]. To ascertain content validity and alignment with clinical realities, item selection and wording were refined through iterative consensus-building discussions among faculty members involved in professional health education.

**Table 1.** Questionnaire content, answer format, and options

| Items | Answer(choice) |
| --- | --- |
| Q1. What is your age? | Select from the following options (20s, 30s, 40s, 50s or more) |
| Q2. What is your sex? | Select from the following options (Male, female, not answering) |
| Q3. Years since obtaining your national qualification (years) | Enter a number |
| Q4. Have you ever had opportunities to learn how to operate mechanical ventilators or care for ventilated patients? | Select from the following options (multiple answers possible)<br>In-person lectures, online seminars, video-on-demand (VOD) contents, simulation-based training, bedside or clinical workplace instruction, No prior opportunities |
| Q5. How familiar are you with the following terms related to MV? | Self-rated understanding of eight key MV terms (FiO<sub>2</sub>, PEEP, Pressure support, CPAP, Assist/Control, Airway pressure, Tidal volume, EtCO<sub>2</sub>, SBT, RST.) assessed on a 4-point scale (1 = Never heard of it, 2 = have heard of it but do not know details, 3 = understand the content, 4 = Can explain to others). |
| Q6. Have you had experience in managing mechanical ventilators or providing care for ventilated patients? | Select from the following options<br>Yer, No |
| Q7. How likely is it that you will be involved in managing mechanical ventilators or providing care for ventilated patients in the future? | 5-point Likert Scale<br>(1: Not at all likely – 5: Very likely) |
| Q8. How confident are you in your ability to engage in tasks related to mechanical ventilation now? | 5-point Likert Scale<br>(1: Not at all confident – 5: Very confident) |
| Q9. How interested would you be in attending a mechanical ventilation training program? | 5-point Likert Scale<br>(1: Not at all willing – 5: Very willing) |
| Q10. If you were to receive education on mechanical ventilation, which formats would you prefer? | Select from the following options (multiple answers possible)<br>In-person lectures, online seminars, VOD contents, simulation-based training, bedside or clinical workplace instruction |
| Q11. How strongly do you wish to learn the following aspects of mechanical ventilation management? | Self-rated willingness to learn for 28 key items related to MV, assessed on 5-point Likert Scale (1: Not at all willing – 5: Very willing)<br><Background knowledge><br>1. Able to understand the basic mechanisms of respiration (anatomy and physiology)/ 2. Able to recall commonly used medications in patients receiving MV/ 3. Able to understand the pharmacological effects of medications commonly used in ventilated patients/ 4. Able to identify potential complications associated with MV/ 5. Able to understand the operational principles of a mechanical ventilator/ 6. Able to understand the different modes of MV<br><Interprofessional collaboration><br>7. Able to understand the role of own profession in MV management/ 8. Able to understand the roles of other professions in MV management/ 9. Able to communicate effectively with other professions regarding MV management<br><Initiation of mechanical ventilation><br>10. Able to understand the goals of MV/ 11. Able to understand the indications for MV/ 12. Able to prepare the necessary equipment and accessories for initiating MV/ 13. Able to recall the key ventilator settings required for initiation<br><MV management><br>14. Able to adjust the ventilator to optimal settings/ 15. Able to understand key observation points for patients on MV/ 16. Able to implement measures to prevent complications of MV/ 17. Able to understand the key items to check on an operating ventilator/ 18. Able to interpret ventilator graphic monitoring displays/ 19. Able to understand ventilator alarm messages/ 20. Able to identify the cause of ventilator alarms/ 21. Able to respond appropriately to the cause of ventilator alarms/ 22. Able to troubleshoot ventilator malfunctions<br><Weaning from mechanical ventilation><br>23. Able to understand the goals of MV management/ 24. Able to understand our institution's ventilator weaning protocol/ 25. Able to understand the risk factors for reintubation/26. Able to assess the risk of reintubation in patients/ 27. Able to prepare the necessary equipment and accessories for ventilator weaning prior to extubating/ 28. Able to understand key observation points after ventilator weaning |
| Q12. What factors do you think facilitate your learning of MV management? | Open-ended response |
| Q13. What factors do you think hinder your learning of MV management? | Open-ended response |

Abbreviation: FiO<sub>2</sub>, Fraction of inspiratory oxygen; PEEP, Positive End-Expiratory Pressure; CPAP, Continuous Positive Airway Pressure; EtCO<sub>2</sub>, end-tidal carbon dioxide; SBT, Spontaneous breathing trial; RST, Respiratory support team; MV, Mechanical Ventilation

### 3. Statistical Analysis

Continuous variables were presented as means and ranges, and categorical variables as frequencies and percentages. Wilcoxon rank-sum and chi-square tests were used to compare groups, with significance set at p < 0.05. The open-ended responses were analysed using content analysis. Coding was conducted independently by two researchers and finalised through consensus. JMP Pro 18.1.1 (JMP Statistical Discovery LLC., Cary, NC, USA) was used for the statistical analysis.

### 4. Ethical Considerations

This study used personal data from health care professionals and was approved by the Ethics Committee of Chiba University Graduate School of Medicine (approval no. M10760). Written informed consent was obtained from all participants. All data were anonymised, and privacy was protected according to Japan’s data protection laws.

## Results

### 1. Participant Characteristics

Table 2 shows participants characteristics. A total of 119 healthcare professionals responded to the survey, including nurses (n=50, 42.0%), medical doctors (n=43, 36.1%), clinical engineers (n=14, 11.8%), pharmacists (n=8, 6.7%), and physical therapists (n=4, 3.4%). The most represented age group was individuals in their 30s (40.3%), with a mean of 11.7 ± 8.3 years since licensure. Overall, 80.6% reported clinical experience with MV, and 84.0% had prior educational exposure, primarily through on-the-job training (67.2%) and in-person lectures (47.9%).

**Table 2.** Responses to the questionnaire regarding participants’ background and current learning status

| Questionnaire | All<br>(n=119) | MD<br>(n=43) | NS<br>(n=50) | Ph<br>(n=8) | PT<br>(n=4) | CE<br>(n=14) |
| --- | --- | --- | --- | --- | --- | --- |
| Q1. Age |  |  |  |  |  |  |
| 20s, n (%) | 32 (26.8%) | 8 (18.6%) | 14 (28%) | 2 (25%) | 0 (0%) | 8 (57.1%) |
| 30s, n (%) | 48(40.3%) | 23(53.5%) | 18(36%) | 3(37.5%) | 1(25%) | 3(21.4%) |
| 40s, n (%) | 29(24.4%) | 8(18.6%) | 14(28%) | 2(25%) | 3(75%) | 2(14.3%) |
| 50s, n (%) | 10(8.4%) | 4(9.3%) | 4(8.0%) | 1(12.5%) | 0(0%) | 1(7.1%) |
| Q2. Sex, n (Male/Female) | 50/69 | 28/15 | 8/42 | 3/5 | 3/1 | 8/6 |
| Q3. Years since obtaining national qualification (years) | 11.7 ± 8.3 | 11.3 ± 7.9 | 12.0 ± 8.5 | 13.8 ± 9.1 | 17.8 ± 3.9 | 9.0 ± 9.3 |
| Q4. Previous learning opportunities |  |  |  |  |  |  |
| In-person lectures, n (%) | 57(47.9%) | 22(51.2%) | 20(40%) | 0(0%) | 4(100%) | 11(78.6%) |
| Online seminars, n (%) | 25(21.0%) | 9(20.9%) | 5(10%) | 0(0%) | 4(100%) | 7(50%) |
| Video-on-demand (VOD) contents, n (%) | 16(13.4%) | 4(9.3%) | 8(16%) | 0(0%) | 1(25%) | 3(21.4%) |
| Simulation-based training, n (%) | 19(16.0%) | 8(18.6%) | 6(12%) | 0(0%) | 1(25%) | 4(28.6%) |
| Bedside or clinical workplace instruction, n (%) | 80(67.2%) | 35(81.4%) | 31(62%) | 0(0%) | 3(75%) | 11(78.6%) |
| No prior opportunities, n (%) | 19(16.0%) | 5(11.6%) | 6(12%) | 8(100%) | 0(0%) | 0(0%) |
| Q5. Understanding of the term |  |  |  |  |  |  |
| Fraction of inspiratory oxygen (FiO2) | 3.4 ± 0.8 | 3.7 ± 0.4 | 3.1 ± 0.9 | 2.1 ± 1.0 | 4 | 3.6 ± 0.5 |
| Positive End-Expiratory Pressure (PEEP) | 3.3 ± 0.8 | 3.7 ± 0.5 | 3.1 ± 0.8 | 1.6 ± 0.5 | 4 | 3.5 ± 0.5 |
| Pressure Support | 3.2 ± 1.0 | 3.6 ± 0.5 | 2.9 ± 1.0 | 1.4 ± 0.5 | 4 | 3.4 ± 0.5 |
| CPAP (mode) | 3.3 ± 0.8 | 3.6 ± 0.6 | 3.1 ± 0.8 | 2.3 ± 0.7 | 4 | 3.5 ± 0.5 |
| Assist/Control | 2.9 ± 1.1 | 3.4 ± 0.7 | 2.5 ± 1.0 | 1.1 ± 0.4 | 2.8 ± 1.5 | 3.4 ± 0.5 |
| Airway pressure | 3.2 ± 0.9 | 3.6 ± 0.6 | 3.0 ± 0.8 | 1.5 ± 0.5 | 4 | 3.4 ± 0.5 |
| Tidal volume | 3.0 ± 1.1 | 3.6 ± 0.7 | 2.5 ± 1.2 | 1.1 ± 0.4 | 4 | 3.5 ± 0.5 |
| end-tidal carbon dioxide (EtCO2) | 3.0 ± 1.1 | 3.6 ± 0.5 | 2.6 ± 1.1 | 1.3 ± 0.5 | 4 | 3.4 ± 0.5 |
| Spontaneous breathing trial (SBT) | 2.5 ± 1.0 | 3.2 ± 0.7 | 2.1 ± 1.0 | 1.4 ± 0.7 | 3.8 ± 0.5 | 2.4 ± 0.9 |
| Respiratory support team (RST) | 2.3 ± 1.0 | 2.8 ± 0.9 | 1.8 ± 0.8 | 1.8 ± 0.7 | 3.5 ± 1.0 | 2.3 ± 1.0 |
| Q6. Experience in managing MV |  |  |  |  |  |  |
| (Yes / No; percentage answering "Yes") | 96/23 (80.6%) | 41/2 (95.3%) | 36/14 (72.0%) | 1/7 (12.5%) | 4/0 (100%) | 14/0 (100%) |
| Q7. Future involvement in MV care | 3.6±1.4 | 3.8±1.3 | 3.4±1.4 | 2.0±1.1 | 4.5±1.0 | 4.1±1.0 |
| Q8. Self-confidence in MV management | 2.4 ± 1.3 | 2.9 ± 1.2 | 2.1 ± 1.2 | 1.1 ± 0.4 | 3.0 ± 0.8 | 2.7 ± 1.0 |
| Q9. Willingness to attend MV training | 3.9 ± 1.1 | 4.1 ± 1.0 | 3.7 ± 1.1 | 3.1 ± 1.2 | 4.0 ± 1.2 | 4.7 ± 0.6 |
| Q10. Preferred formats for MV education |  |  |  |  |  |  |
| In-person lectures | 53 (44.5%) | 20 (46.5%) | 20 (40.0%) | 3 (37.5%) | 3 (75.0%) | 7 (50.0%) |
| Online seminars | 56 (47.1%) | 18 (41.9%) | 22 (44.0%) | 5 (62.5%) | 3 (75.0%) | 8 (57.1%) |
| Video-on-demand (VOD) contents | 63 (52.9%) | 19 (44.2%) | 26 (52.0%) | 6 (75.0%) | 4 (100%) | 8 (57.1%) |
| Simulation-based training | 73 (61.3%) | 34 (79.1%) | 26 (52.0%) | 2 (25.0%) | 2 (50.0%) | 9 (64.3%) |
| Bedside or clinical workplace instruction | 54 (45.4%) | 27 (62.8%) | 27 (54.0%) | 3 (37.5%) | 1 (25.0%) | 6 (42.9%) |

Abbreviation: MD, Medical doctor; NS, Nurse; Ph, Pharmacist; PT, Physical Therapist; CE, Clinical Engineer, MV. Mechanical Ventilation

The self-rated understanding of MV-related terminology was generally higher among medical doctors, clinical engineers, and physical therapists. Significant differences (p<0.01) were identified across professions in self-rated understanding of MV terminology, primarily driven by lower scores among pharmacists. Except for the item ‘Respiratory Support Team’, participants with clinical experience in MV showed significantly greater understanding of all terminology items.

The average confidence level in managing MV was modest (2.4 ± 1.3 on a 5-point scale), while motivation to participate in MV education was relatively high (3.9 ± 1.1). Participants with clinical experience in MV reported significantly higher motivation compared to those without such experience (4.1 ± 1.0 vs. 3.2 ± 1.0, p < 0.01). Furthermore, the number of years since licensure showed a significant negative correlation with motivation (p = 0.01).

Regarding preferred learning modalities, simulation-based education was the most favoured format (61.3%), followed by bedside teaching and on-demand sessions (52.9%). Profession-based trends were observed; medical doctors and clinical engineers revealed a stronger preference for simulations, whereas pharmacists and physical therapists preferred on-demand learning.

### 2. Educational Needs

As summarised in Table 3, the mean score for all 28 learning items exceeded 4.0 on a 5-point scale, indicating consistently high interest across all domains, including foundational knowledge, initiation, clinical management, ventilator weaning, and interprofessional collaboration.

**Table 3.** Self-rated willingness to learn for 28 key items related to mechanical ventilation (1: Not at all willing – 5: Very willing)

| Questionnaire | All<br>(n=119) | MD<br>(n=43) | NS<br>(n=50) | RPh<br>(n=8) | PT<br>(n=4) | CE<br>(n=14) | p value |
| --- | --- | --- | --- | --- | --- | --- | --- |
| <Background knowledge> |  |  |  |  |  |  |  |
| 1. understand the basic mechanisms of respiration | 4.1 ± 0.8 | 4.0 ± 0.9 | 4.1 ± 0.9 | 4.3 ± 0.7 | 4.5 ± 0.6 | 4.4 ± 0.8 | 0.77 |
| 2. recall commonly used medications in MV | 4.2 ± 0.9 | 4.2 ± 0.9 | 4.1 ± 1.0 | 4.9 ± 0.4 | 4.5 ± 0.6 | 4.2 ± 0.9 | 0.21 |
| 3. understand the pharmacological effects of medication | 4.2 ± 0.9 | 4.1 ± 0.9 | 4.1 ± 0.9 | 4.9 ± 0.4 | 4.5 ± 0.6 | 4.1 ± 1.0 | 0.18 |
| 4. identify potential complications associated with MV | 4.3 ± 0.9 | 4.2 ± 0.9 | 4.2 ± 0.9 | 4.4 ± 0.5 | 4.5 ± 0.6 | 4.8 ± 0.4 | 0.27 |
| 5. understand the operational principles of MV | 4.2 ± 0.9 | 4.2 ± 0.7 | 4.2 ± 1.0 | 4.4 ± 0.7 | 4.0 ± 1.4 | 4.4 ± 0.9 | 0.85 |
| 6. understand the different modes of MV | 4.3 ± 0.9 | 4.4 ± 0.7 | 4.3 ± 1.0 | 3.6 ± 1.2 | 4.5 ± 0.6 | 4.6 ± 0.6 | 0.10 |
| <Interprofessional collaboration> |  |  |  |  |  |  |  |
| 7. understand the role of own profession | 4.2 ± 0.9 | 4.3 ± 0.9 | 4.1 ± 1.0 | 4.1 ± 0.8 | 4.3 ± 1.0 | 4.4 ± 0.6 | 0.80 |
| 8. understand the roles of other professions | 4.2 ± 0.9 | 4.3 ± 1.0 | 4.1 ± 0.9 | 4.4 ± 0.7 | 4.3 ± 1.0 | 4.4 ± 0.7 | 0.68 |
| 9. communicate effectively with other professions | 4.3 ± 0.9 | 4.4 ± 0.9 | 4.1 ± 1.0 | 4.4 ± 0.7 | 4.3 ± 1.0 | 4.4 ± 0.7 | 0.53 |
| <Initiation of mechanical ventilation> |  |  |  |  |  |  |  |
| 10. understand the goals of MV | 4.2 ± 0.9 | 4.1 ± 0.9 | 4.0 ± 1.0 | 4.5 ± 0.8 | 4.3 ± 1.0 | 4.5 ± 0.7 | 0.39 |
| 11. understand the indications for MV | 4.2 ± 0.9 | 4.2 ± 0.9 | 4.1 ± 0.9 | 4.1 ± 1.0 | 4.3 ± 1.0 | 4.6 ± 0.6 | 0.57 |
| 12. prepare the necessary equipment for initiation | 4.1 ± 1.0 | 4.4 ± 0.6 | 4.2 ± 1.0 | 2.4 ± 1.1 | 3.8 ± 1.3 | 4.3 ± 0.9 | <0.01 |
| 13. recall the key ventilator settings for initiation | 4.2 ± 1.0 | 4.4 ± 0.9 | 4.2 ± 0.9 | 2.8 ± 1.5 | 4.3 ± 1.0 | 4.5 ± 0.7 | <0.01 |
| <Mechanical ventilation management> |  |  |  |  |  |  |  |
| 14. adjust the ventilator to optimal settings | 4.1 ± 1.1 | 4.4 ± 0.8 | 4.1 ± 1.0 | 1.9 ± 0.6 | 4.3 ± 1.0 | 4.3 ± 0.9 | <0.01 |
| 15. understand key observation points for patients on MV | 4.3 ± 0.8 | 4.5 ± 0.7 | 4.3 ± 1.0 | 4.1 ± 0.8 | 4.3 ± 1.0 | 4.4 ± 0.9 | 0.73 |
| 16. implement measures to prevent complications of MV | 4.3 ± 0.9 | 4.4 ± 0.8 | 4.3 ± 1.0 | 3.4 ± 1.1 | 4.3 ± 1.0 | 4.1 ± 0.9 | 0.048 |
| 17. understand the key items to check on MV | 4.3 ± 0.9 | 4.5 ± 0.7 | 4.3 ± 0.9 | 3.0 ± 1.3 | 4.3 ± 1.0 | 4.6 ± 0.5 | <0.01 |
| 18. interpret ventilator graphic monitoring displays | 4.3 ± 0.9 | 4.4 ± 0.8 | 4.2 ± 1.0 | 3.6 ± 1.3 | 4.5 ± 0.6 | 4.6 ± 0.6 | 0.11 |
| 19. understand ventilator alarm messages | 4.3 ± 1.0 | 4.4 ± 0.9 | 4.2 ± 1.0 | 3.3 ± 1.4 | 4.3 ± 1.0 | 4.5 ± 0.7 | 0.04 |
| 20. identify the cause of ventilator alarms | 4.3 ± 1.0 | 4.5 ± 0.8 | 4.3 ± 1.0 | 2.5 ± 0.9 | 4.5 ± 0.6 | 4.4 ± 0.6 | <0.01 |
| 21. respond appropriately to the cause of ventilator alarms | 4.3 ± 1.0 | 4.6 ± 0.7 | 4.3 ± 0.9 | 2.1 ± 0.8 | 4.3 ± 1.0 | 4.5 ± 0.7 | <0.01 |
| 22. troubleshoot ventilator malfunctions | 4.3 ± 1.0 | 4.6 ± 0.7 | 4.3 ± 1.0 | 2.3 ± 1.0 | 4.3 ± 1.0 | 4.5 ± 0.7 | <0.01 |
| <Weaning from mechanical ventilation> |  |  |  |  |  |  |  |
| 23. understand the goals of mechanical... | 4.3 ± 0.8 | 4.4 ± 0.8 | 4.2 ± 0.9 | 4.5 ± 0.8 | 4.3 ± 1.0 | 4.1 ± 0.8 | 0.54 |
| 24. understand an institution's ventilator weaning protocol | 4.2 ± 0.9 | 4.5 ± 0.6 | 4.1 ± 1.0 | 3.5 ± 1.3 | 4.3 ± 1.0 | 4.1 ± 0.9 | 0.04 |
| 25. understand the risk factors for reintubation | 4.3 ± 0.9 | 4.4 ± 0.8 | 4.1 ± 1.0 | 3.8 ± 1.2 | 4.5 ± 0.6 | 4.3 ± 0.7 | 0.22 |
| 26. assess the risk of reintubation | 4.1 ± 1.0 | 4.4 ± 0.8 | 4.0 ± 1.0 | 2.9 ± 1.0 | 4.8 ± 0.5 | 4.1 ± 0.9 | <0.01 |
| 27. prepare the necessary equipment for weaning | 4.0 ± 1.0 | 4.5 ± 0.7 | 4.0 ± 1.0 | 2.0 ± 0.8 | 3.8 ± 1.3 | 3.9 ± 0.9 | <0.01 |
| 28. understand key observation points after weaning | 4.2 ± 0.9 | 4.4 ± 0.8 | 4.2 ± 0.9 | 3.6 ± 1.1 | 4.5 ± 0.6 | 4.1 ± 0.8 | 0.19 |

abbreviation: MV, Mechanical ventilation; MD, Medical doctor; NS, Nurse; Ph, Pharmacist; PT, Physical Therapist; CE, Clinical Engineer

Although learning needs were uniformly high, profession-based differences emerged in specific domains. Notable variation was observed in areas such as equipment preparation (e.g. ‘Able to prepare the necessary equipment and accessories for initiating mechanical ventilation’, p<0.01), alarm management (e.g. ‘Able to identify the cause of ventilator alarms’, p<0.01), reintubation risk assessment (e.g. ‘Able to assess the risk of reintubation in patients’, p<0.01), and prevention of complications (e.g. ‘Able to implement measures to prevent complications of mechanical ventilation’, p<0.05). Conversely, no significant differences were found between the professions in terms of foundational knowledge or collaboration, indicating shared baseline educational priorities.

### 3. Facilitators and Barriers

As shown in Table 4, the content analysis of free-text responses identified five primary facilitators of MV learning: (1) opportunities for experiential learning (especially bedside involvement), (2) workplace conditions enabling learning, (3) access to structured educational resources (e.g. in-house seminars and e-learning), (4) intrinsic motivation and perceived relevance, and (5) organisational support and supervisory guidance. These facilitators were commonly cited across all professional groups, and no statistically significant differences were observed between the professions.

**Table 4.** Facilitators and barriers to learning mechanical ventilation

| Category | Key words | n | Quotes |
| --- | --- | --- | --- |
| **Facilitators of Learning** |  |  |  |
| Experiential Learning through Clinical Exposure | Clinical practice, Patient management, Intensive care | 47 | To care for patients who are actually on mechanical ventilation. |
| Workplace Conditions Enabling Learning | Securing time, workload, | 32 | Having an appropriate workload |
| Structured Access to Educational Opportunities | e-learning, simulation, workshop | 24 | If announcements about training sessions or workshops are made, I would actively consider participating. |
| Intrinsic Motivation and Perceived Relevance | Necessity, Certification acquisition, Interest | 17 | Recognizing the necessity of ventilator management is the first step toward proactive learning. |
| Organizational Support and Supervisory Guidance | Competent instructors, Feedback | 15 | Receiving timely feedback during the care of patients on mechanical ventilation. |
| None |  | 3 |  |
| **Barriers to Learning** |  |  |  |
| Excessive Workload and Time Constraints | Fatigue, Heavy workload, Aversion to after-hours training | 50 | The demanding nature of daily clinical duties leaves little room for engaging in learning activities. |
| Limited Opportunities for Hands-on Practice | Lack of exposure, Department rotation | 30 | A work environment where mechanical ventilation cases are not encountered. |
| Lack of Motivation and Goal Orientation | Priority, Sense of difficulty, Professional roles | 16 | I don't understand how learning about this would benefit my professional role. |
| Limited Access to Educational Resources and Opportunities | Learning opportunity, Practical workshop | 15 | Skills cannot be effectively acquired without hands-on learning using actual ventilator equipment. |
| Inadequate Supervisory and Peer Support Systems | Financial burden, Instructor, Structured education | 8 | My workplace does not have a supervisor with sufficient expertise in ventilator management. |
| None |  | 11 |  |

Barriers to learning were also prominent, including excessive workload, time constraints, insufficient clinical exposure to ventilated patients, low individual motivation, limited access to training opportunities, and inadequate supervision or feedback. Table 5 presents the profession-wise distribution of learning facilitators and barriers. Workload-related challenges demonstrated significant variation across professions (p = 0.01), with notably high proportions among physical therapists (4 out of 4, 100%) and nurses (27 out of 50, 54.0%). Similarly, lack of motivation as a barrier differed significantly across the groups (p = 0.02), with pharmacists reporting the highest frequency (3 out of 8, 37.5%).

**Table 5.** Facilitators and barriers to mechanical ventilation education by professional category

| Category | All<br>(n=119) | MD<br>(n=43) | NS<br>(n=50) | Ph<br>(n=8) | PT<br>(n=4) | CE<br>(n=14) | p value |
| --- | --- | --- | --- | --- | --- | --- | --- |
| **Facilitators of Learning** |  |  |  |  |  |  |  |
| Experiential Learning through Clinical Exposure | 47(39.5%) | 17(39.5%) | 23(46%) | 3(37.5%) | 1(25%) | 3(21.4%) | 0.53 |
| Workplace Conditions Enabling Learning | 32(26.9%) | 12(27.9%) | 13(26%) | 2(25%) | 2(50%) | 3(21.4%) | 0.85 |
| Structured Access to Educational Opportunities | 24(20.2%) | 10(23.3%) | 7(14%) | 2(25%) | 1(25%) | 4(28.6%) | 0.69 |
| Intrinsic Motivation and Perceived Relevance | 17(14.3%) | 6(14.0%) | 10(20%) | 0(0%) | 0(0%) | 1(7.1%) | 0.42 |
| Organizational Support and Supervisory Guidance | 15(12.6%) | 6(14.0%) | 4(8.0%) | 0(0%) | 1(25%) | 4(28.6%) | 0.2 |
| None | 3(2.5%) | 2(4.7%) | 1(2.0%) | 0(0%) | 0(0%) | 0(0%) | 0.82 |
| **Barriers to Learning** |  |  |  |  |  |  |  |
| Excessive Workload and Time Constraints | 50(42.4%) | 13(30.2%) | 27(54%) | 3(37.5%) | 4(100%) | 3(21.4%) | **0.01 \*** |
| Limited Opportunities for Hands-on Practice | 30(25.4%) | 16(37.2%) | 11(22%) | 1(12.5%) | 0(0%) | 2(14.3%) | 0.18 |
| Lack of Motivation and Goal Orientation | 16(13.6%) | 6(14.0%) | 2(4.0%) | 3(37.5%) | 1(25%) | 4(28.6%) | **0.02 \*** |
| Limited Access to Educational Resources and Opportunities | 15(12.7%) | 8(18.6%) | 4(8.0%) | 0(0%) | 0(0%) | 3(21.4%) | 0.29 |
| Inadequate Supervisory and Peer Support Systems | 8(6.8%) | 2(4.7%) | 4(8.0%) | 1(12.5%) | 0(0%) | 1(7.1%) | 0.85 |
| None | 11(9.3%) | 5(11.6%) | 4(8.0%) | 0(0%) | 0(0%) | 2(14.3%) | 0.75 |

abbreviation: MD, Medical doctor; NS, Nurse; Ph, Pharmacist; PT, Physical Therapist; CE, Clinical Engineer

## Discussion

This study identified both common and profession-specific learning needs in MV management, providing clear direction for the development of effective IPE programs aimed at enhancing preparedness for future health emergencies. Our findings underscore both shared and profession-specific aspects of MV education, offering practical guidance for developing structured team-based educational programs.

The consistently high motivation and educational interest across all professional groups indicate a strong collective awareness of the importance of MV competencies. This aligns with the core premise of IPE, which emphasises the development of collaborative competencies through shared learning experiences [19]. The relatively high motivation observed among those with clinical experience in MV indicates that experiential exposure reinforces the perceived relevance and urgency of MV-related knowledge, a concept consistent with the adult learning theory and experiential learning frameworks [20] [21].

Our findings revealed significant profession-based differences in baseline understanding and preferred learning formats. Simulation-based training aligns with strong evidence that it significantly improves MV-related procedural skills, confidence, and knowledge retention among trainees [22] [23]. This may partly explain the preferences observed among medical doctors and clinical engineers who frequently engage in ventilator operations and acute decision-making. Conversely, pharmacists and physical therapists, roles that typically do not involve direct ventilator operations or emergent clinical decision-making, may prefer on-demand asynchronous e-learning formats. This preference is consistent with the finding that pharmacists value flexible, self-paced learning in continuing education contexts [24]. Similarly, in physiotherapy education, digitally enhanced or asynchronous learning modalities are increasingly being welcomed owing to their practical accessibility, cost-effectiveness, and ability to support self-directed engagement [25].

Hence, we propose a two-tiered instructional model that integrates a common core curriculum focusing on shared foundational knowledge and interprofessional collaboration with profession-specific modules tailored to each discipline’s unique clinical roles and learning preferences [26] [27]. For example, simulation-based scenarios could be emphasised for medical doctors and clinical engineers, whereas pharmacists and physical therapists could benefit from asynchronous e-learning modules that underscore pharmacological considerations or ventilator-related rehabilitation strategies.

Importantly, several barriers to learning that disproportionately affected certain professions were identified. For example, physical therapists and nurses frequently cited workload as a barrier, whereas pharmacists reported low motivation. Although subgroup sizes were limited in some cases, these findings suggest profession-specific patterns in perceived barriers. These findings highlight the need for organisational strategies to reduce learning-related burdens and foster a culture that values continuous interprofessional development [28]. The proposed model aligns with the principles of high-functioning healthcare teams, including role clarity, mutual support, and situational awareness. These principles are central to Team Strategies and Tools to Enhance Performance and Patient Safety (TeamSTEPPS^™^), which emphasise structured communication and interprofessional collaboration [29]. Our qualitative findings support these principles: participants emphasised the importance of workplace conditions, organisational support, and structured educational opportunities, factors known to enhance interprofessional team performance [30] [31].

## Limitations

This single-centre survey may have limited the generalisability of the results. Regional and institutional factors may also affect applicability. The study evaluated perceptions rather than competencies, and the respondents were likely to include a higher proportion of experienced personnel. The free-text analysis was limited by variations in response details, and qualitative interviews may have provided richer insights.

## Conclusion

This study highlights the strong shared motivation among healthcare professionals to engage in MV education alongside distinct profession-specific learning needs. While foundational knowledge and inter-professional collaboration are commonly valued, priorities and barriers vary across roles. These findings support the development of IPE programs that combine a shared core curriculum with tailored modules to address discipline-specific competencies. To enhance individual expertise and team-based readiness, future efforts should emphasise on context-sensitive, team-oriented educational strategies supported by longitudinal evaluations to assess their impact on practice and patient outcomes.

## Data Availability

All relevant data will be available from the Dryad Digital Repository upon acceptance.

## Acknowledgements

This work was supported by JSPS KAKENHI (grant number 24K13283).

